# The genetic relationships between brain structure and schizophrenia

**DOI:** 10.1101/2023.03.13.23287137

**Authors:** Eva-Maria Stauffer, Richard A.I. Bethlehem, Lena Dorfschmidt, Hyejung Won, Varun Warrier, Edward T. Bullmore

## Abstract

Recent studies suggest shared genetic effects on both schizophrenia and brain structure, but it has been challenging to specify which genes mediate this pleiotropic association. We accessed genome-wide association data on schizophrenia (N=69,369 cases; 236,642 controls), and on three magnetic resonance imaging (MRI) metrics (surface area, cortical thickness, neurite density index) measured at 180 cortical areas (N=36,843). Using Hi-C-coupled MAGMA, we identified 61 genes that were significantly associated with both schizophrenia and one or more MRI metrics. Whole genome analysis demonstrated significant genetic covariation between schizophrenia and area or thickness of most cortical regions. Genetic similarity between cortical areas was strongly coupled to covariance of their MRI metrics, and genetic covariation between schizophrenia and cortical regional phenotypes was greatest in the hubs of the corresponding structural covariance network. Three genomic regions, on chromosomes 3p21, 17q21 and 11p11, were enriched for neurodevelopmental processes and consistently implicated in these pleiotropic associations between schizophrenia and cortical network organization.

## Introduction

Anatomical variation in brain cortical structure is associated with cognitive and emotional processes as well as with psychiatric disorders, such as schizophrenia [1]. Recent large scale, genome-wide association studies (GWAS) showed that cortical morphology is heritable [2, 3]. Furthermore, the number of genetic loci associated with variation in human brain phenotypes, often derived from magnetic resonance imaging (MRI) data, is increasing as the scale of available cohorts with both genetic and MRI data has increased in recent years [2, 3]. Growing awareness of the genetic architecture of the human brain raises the question whether brain structural variation in the population is associated with genes that are also significantly associated with schizophrenia [4, 5, 6, 7]. It is a central expectation of many biological theories of schizophrenia that genetic effects on schizophrenia are mechanistically or proximally mediated by the effects of the same genes on brain structure and function [8]. Here we aimed to identify genes that are associated with both brain MRI phenotypes and schizophrenia. We reasoned that identification of such pleiotropic genes would be consistent with prior theories that genetic variants encode risk for schizophrenia by affecting intermediate endo-phenotypes of brain structure. Conversely, the absence of any such gene set would refute the hypothesis that genetic risk for schizophrenia is mediated by brain structure [9, 10]. There is limited evidence from previous studies that there might indeed be a genetic overlap between schizophrenia and brain structure [4, 5, 6, 7]; but it has proven challenging to specify which individual genes are most strongly pleiotropic for three key reasons: (i) prior focus on macro-structural MRI measurements, (ii) regionally varied genetic associations with brain structure, and (iii) methodological considerations for gene identification.

First, the structure of the human cortex can be quantified in terms of a large number of brain metrics estimated from diverse MRI or diffusion-weighted imaging (DWI) measurements. Most genetic-MRI studies have used T1-weighted data to estimate macro-structural phenotypes such as surface area (SA), cortical thickness (CT), and volume [2], each estimated at multiple cortical areas or globally. However, micro-structural MRI metrics are increasingly recognised to provide important additional information about cortical myelination, lamination and other tissue properties, e.g., the density of axons and dendrites measured by neurite density index (NDI) [11]. Both macro- and micro-structural MRI metrics are heritable and can be organised into latent dimensions of shared and distinctive genetic effects [3]. We therefore considered it important to measure genetic associations separately for three minimally correlated MRI phenotypes (CT, SA, NDI) that developmentally emerge from distinct cellular mechanisms [3], and to test the hypothesis (H1) that specific genes can have effects on schizophrenia and brain structure that are either specific to a particular MRI metric, or shared across metrics.

Second, genetic associations with brain structure can be measured globally or locally at each of hundreds of cortical areas defined by a standard cortical parcellation scheme. Recent GWAS data on regional MRI phenotypes have shown that single nucleotide polymorphisms (SNPs) often have specific regional effects that are not shared globally, so we focused on regional MRI phenotypes [2, 3, 12]. Furthermore, given that the cortex is organised as a complex network, it is important to consider the relationship between genetic variation and brain network phenotypes. For example, a structural covariance network [13], estimated from the inter-regional covariance of a structural MRI metric in a group of participants, has non-random topological features, such as highly connected hub nodes (brain regions), that are strongly heritable in twin studies [14] and often the locus of atypical brain structure in case-control studies of schizophrenia [15, 16]. Following Cheverud’s conjecture [17], that phenotypic correlations mirror genetic correlations [17], we predicted that structural covariance and genetic similarity between cortical areas should be strongly coupled. We also tested the hypothesis (H2) that the strength of genetic overlap between schizophrenia and regional cortical brain structure is related to the topological properties of each cortical node in the structural covariance network or connectome derived from each MRI metric.

Third, there are challenges in measuring convergence of genetic effects on brain structural and clinical phenotypes. Previous studies addressed this question by performing genetic correlation analysis or by relating polygenic scores (PGS) for schizophrenia to brain structural phenotypes [2, 6, 18, 19]. However, genetic correlations and PGS analyses are unable to pinpoint specific genes or biological processes that are implicated in both brain structure and schizophrenia, as they are composite measures of a large number of SNPs across the genome [8]. To identify specific genes and shared biological mechanisms, the genetic relationship between brain structure and schizophrenia can be more directly investigated by mapping SNPs to genes. We therefore accessed recently published GWAS statistics for three MRI metrics (CT, SA and NDI), each measured at 180 regions in N = 36,843 scans from the UK Biobank and the ABCD cohort [3]. We also accessed summary statistics from the largest and most recent GWAS study of schizophrenia (N = 69,369 individuals with a schizophrenia diagnosis and N = 236,642 individuals without a schizophrenia diagnosis) [20]. We mapped each set of GWAS statistics to individual genes using H-MAGMA [21, 22], which takes into account that non-coding SNPs can regulate distal genes via chromatin interaction profiles (measured by high-throughput chromosome conformation capture, Hi-C). Additionally, H-MAGMA allows developmental stage-specific gene mapping by integrating chromatin interaction profiles from fetal brain (gestation weeks 17–18) or adult brain (age 36-64 years) Hi-C datasets [21, 22].

On this basis, we identified and characterised genes significantly associated with one or more of three regional MRI metrics (SA, CT and NDI) [3] and the subset of MRI-related genes that were also significantly associated with schizophrenia. We used partial least squares (PLS), a dimension-reducing multivariate technique, for whole genome analysis and cortical mapping of genetic covariation between schizophrenia and each of the MRI metrics. We investigated the coupling between genetic similarity and structural covariance for each MRI metric and demonstrated that genetic covariation with schizophrenia was highest in cortical areas constituting highly connected hubs of structural covariance networks. Finally, we tested the pleiotropic gene set identified by PLS analysis for positional and functional enrichment.

## Results

### Genetic associations with regional MRI phenotypes

#### Identification of genes associated with MRI metrics

First, we focused on identifying and characterising genes that were significantly associated with each MRI metric. We performed SNP-to-gene mapping for each GWAS using H-MAGMA [21] (Methods). We used H-MAGMA for two reasons; first, compared to gene-mapping methods that only use positional information, H-MAGMA can incorporate tissue-specific and long-range regulatory effects [21]; second, although transcription-based gene-mapping methods (TWAS) can integrate gene regulatory information, currently available fetal brain eQTL datasets do not have large-enough sample sizes, and do not capture trans-eQTL effects well [23, 24]. Using H-MAGMA we were able to map SNPs to a total of 18,640 protein-coding genes for each GWAS.

Across all 180 brain regions we identified 4,222 significant gene-level associations for SA, 773 for CT and 301 for NDI, after accounting for multiple comparisons within each metric (Methods). For each MRI metric, the number of significantly associated genes varied between cortical regions (Fig.1 A). However, most of the genes were significantly associated with multiple cortical regions, suggesting that shared genetic factors influenced variation of each phenotype across the cortex, which is in line with high genetic correlations across regions [2, 3]. Aggregating associated genes across all regions, we identified 318 genes in total for SA (i.e. 4,222 significant gene-region associations represent 318 non-redundant genes), 157 genes for CT and 86 genes for NDI (Table S1).

**Figure 1.**
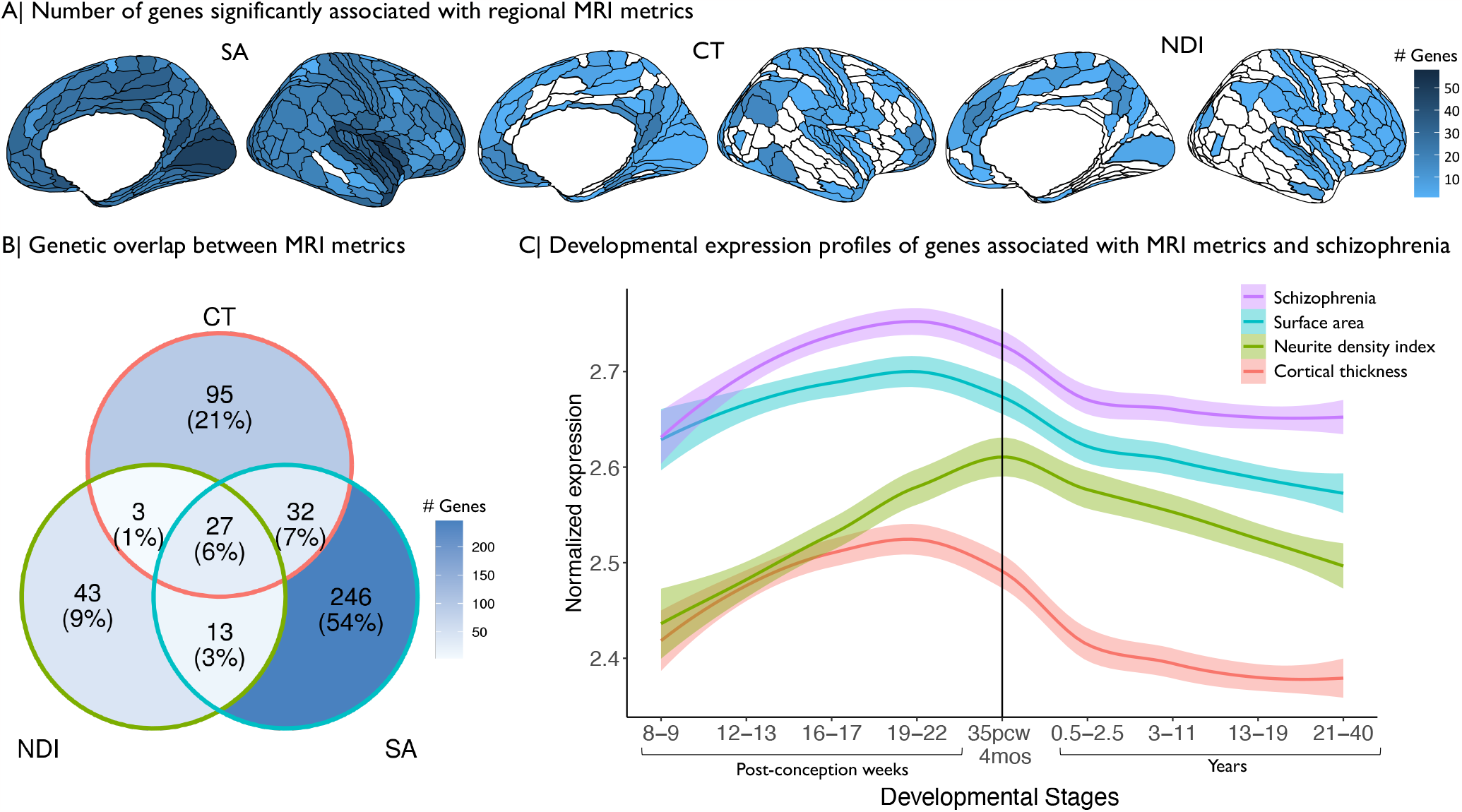
Genetic associations with three MRI metrics of regional brain structure. surface area (SA), cortical thickness (CT) and neurite density index (NDI). (A) Cortical surface maps representing the number of genes significantly associated with variation in each MRI metric at each of 180 cortical areas, from left to right: SA, CT, NDI. Regions without any significant gene associations are shown in white. (B) Venn diagram representing the number of genes that are specifically associated with each MRI metric or generically associated with two or three metrics. The percentages refer to the proportion of all genes associated with one or more MRI metrics represented in each segment of the Venn diagram. (C) Developmental trajectories of average gene expression from 8 PCW to 40 years for the sets of genes significantly associated with each MRI metric or with schizophrenia (SCZ). The vertical line indicates the usual timing of birth.

We found that many of these genes were uniquely associated with one of the three MRI metrics: 78% of the genes associated with SA were associated only with SA (246 out of 318 genes), 61% for CT (95 out of 157 genes), and 63% for NDI (43 out of 86 genes). However, 27 genes were significantly associated with all MRI metrics, including 16 genes within the 17q21.31 region, which contains an inversion polymorphism with a complex linkage disequilibrium (LD) structure [25]. The remaining (11) genes associated with all 3 MRI metrics were located on chromosome 8p23 (seven genes), chromosome 6q25 (three genes) and one gene on chromosome 1p33 (Fig.1 B, Table S2). In short, effects of genetic variation shared across all brain MRI phenotypes were localised at four chromosomal regions.

#### Pathway enrichment and developmental analysis of genes associated with MRI metrics

We found that the set of 318 genes associated with SA across multiple cortical areas was enriched for 34 Gene Ontology (GO) terms, and the set of 157 genes associated with CT was enriched for 12 GO terms (Methods). SA-related genes were enriched in processes related to central nervous system development (SA GO:0007417, *P* = 0.01) and neurogenesis (SA GO:0050767, *P* < 0.05). CT-related genes were enriched for neuron development (CT GO:0048666, *P* = 0.01), neuron projection (CT GO:0031175, *P* = 0.01) and microtubule related processes (CT, GO:0007017, *P* < 0.002). Both SA- and CT-related gene sets were enriched for fundamental biological processes such as cell development (SA GO:0048869 < *P* = 0.01, CT GO:0048468, < *P* = 0.05) (Table S3). The 86 genes associated with NDI were not significantly enriched for any biological pathways.

Using spatiotemporal gene expression data from the PsychEncode database [26], we investigated the developmental expression profiles of gene sets significantly associated with each MRI metric (Methods). Genes associated with CT and SA showed remarkably similar trajectories with peak expression during the mid-gestation period (developmental stage 4, 19-22 post-conception weeks (PCW)) followed by a steep decline of expression post-natally. Expression of genes associated with NDI peaked later, in the peri-natal period (developmental stage 5, 35 PCW - 4 months), and gradually decreased post-natally (Fig.1 C; Table S4). These results highlight mid-to-late fetal stages as a critical window for genetically controlled development of cortical regions and are consistent with findings from global MRI phenotypes [3].

#### Genes associated with schizophrenia and their intersection with MRI-associated genes

We compared the MRI-associated gene sets with genes identified as significant based on H-MAGMA analysis of an independent GWAS of schizophrenia [20]. We found 587 genes were significantly associated with schizophrenia after correction for multiple comparisons (Table S5) (Methods). In line with previous findings [20, 21], this gene set was significantly enriched for 66 GO terms related to neuronal function including nervous system development (GO:0007399, *P* < 0.001), neurogenesis (GO:0022008, *P* < 0.001) and trans-synaptic transmission (GO:0099537, *P* < 0.01) (Table S6). Genes associated with schizophrenia had peak expression during mid-gestation followed by decreasing expression post-natally (Fig.1 B; Table S4), as reported previously [21].

Out of 587 schizophrenia-associated genes, 51 were also associated with SA, 22 with CT and 14 with NDI, representing a significant overlap for each metric (SA *Z* = 16.5, CT *Z* = 8.91, NDI *Z* = 8.14, all *P* < 0.0001 by permutation tests; see Table S7 for details). The genomic region of chromosome 3p21.1 contained several genes pleiotropically associated with both SA and schizophrenia, including *PBRM1, NEK4, GNL3, ITIH4* and *NISCH*. Likewise the genomic region of chromosome 17q21.31 also contained multiple genes that were associated both with schizophrenia and with all 3 MRI metrics, including *NSF, KANSL1, CRHR1, ARHGAP27, LRRC37A, CCDC43, FMNL1, SPPL2C, MAPT, PLEKHM1, STH*, and *LINC002210-CRHR1*.

### Genetic covariation between schizophrenia and regional MRI metrics

Since the number of significantly associated genes varied between cortical regions and MRI metrics, and because the genetic covariation between schizophrenia and brain structure might be influenced by genes that do not reach genome-wide significance, we further investigated the genetic relationship between brain structure and schizophrenia without selecting genes based on a *P*-value threshold.

Using partial least squares regression (PLS) [27] to map the anatomical distribution of genetic covariation between brain regional structure and schizophrenia (Fig. 2 A, Methods), we found that the first PLS component for each MRI metric identified a modest but significant proportion of its genetically determined variation that covaried with genetic risks for schizophrenia: 5.9% for SA, 5.5% for CT and 3% for NDI (all *P* < 0.0001 by permutation tests). PLS1 weights for each brain region, 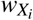, were *Z*-transformed based on bootstrapped standard errors and tested for statistical significance using the false discovery rate (FDR = 5%) to correct for multiple comparisons (180 regional tests). The strength of genetic covariation with schizophrenia differed between MRI metrics, with the greatest mean PLS1 weight for SA 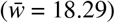, followed by CT 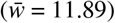 and NDI 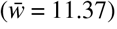.

**Figure 2.**
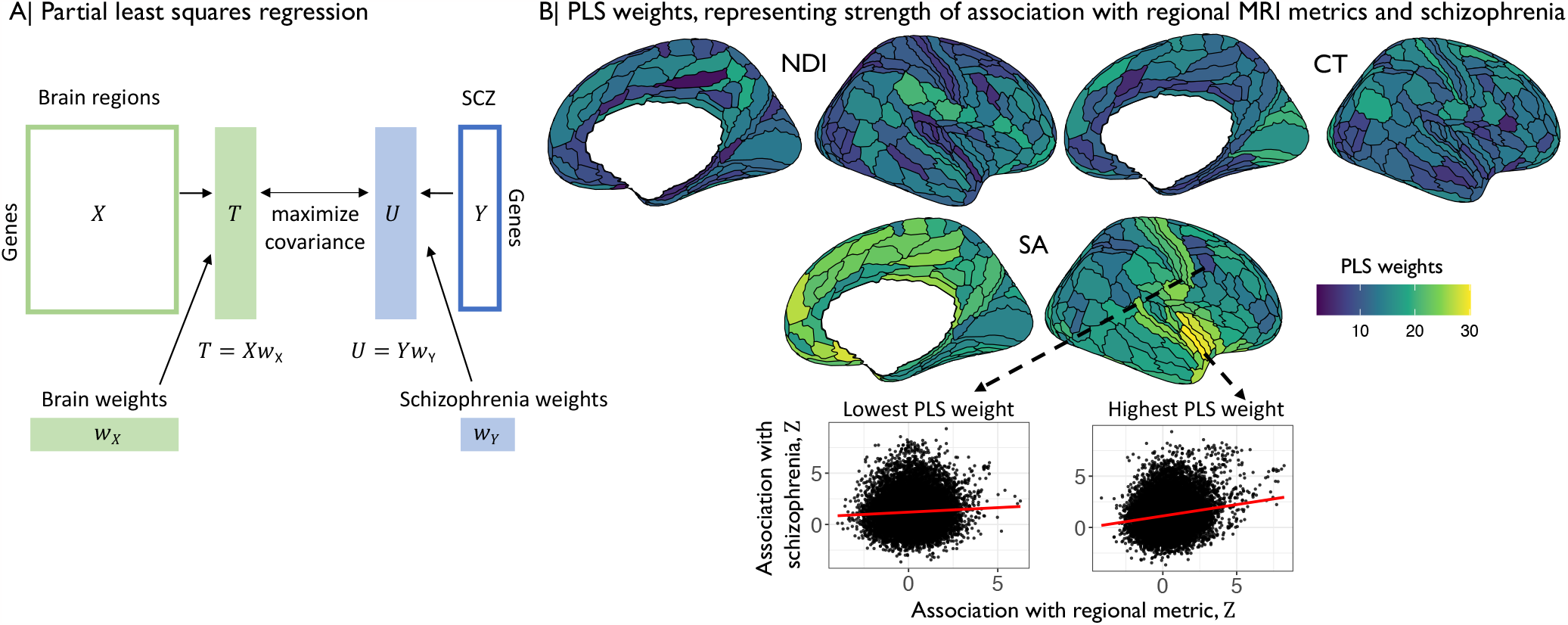
Partial least squares analysis of genetic covariation between regional MRI metrics and schizophrenia. (A) The {1 *×* 18,640 } vector of unthresholded gene association statistics (*Z*-scores) derived by H-MAGMA analysis of the schizophrenia GWAS dataset was designated as the response variable, i.e., the dependent *Y* vector; and the {180 *×* 18,640} matrix of unthresholded gene association *Z*-scores derived by H-MAGMA analysis of each of the MRI GWAS datasets was designated the predictor variable, i.e., the independent *X* matrix. PLS generally finds the set of orthogonal components or weighted functions of *X* that are ranked in terms of their covariance with *Y*. In this case, the first PLS component (PLS1) represented the weighted function of *Z*-scored gene associations with each regional MRI metric that was most strongly correlated with the *Z*-scored gene associations with schizophrenia, over all genes in the genome. More specifically, the PLS1 weights for *X* (brain weights, *w*_*X*_) multiplied by *X* constituted a {1 *×* 18,640} vector of *T* scores (genes weighted by association with brain phenotypes), whereas the PLS1 weights for *Y* (schizophrenia weights *w*_*Y*_) multiplied by *Y* constituted a {1 *×* 18,640} vector of *U* scores (genes weighted by association with schizophrenia), such that the covariance between *T* and *U* scores was maximal over all possible weighted functions of *X* and *Y*. Thus genes with the highest absolute *T* and *U* scores can be regarded as the genes which contribute most strongly to the genetic covariation between schizophrenia and each MRI metric of brain structure [27, 30]. (B) Cortical surface maps of standardised PLS1 weights for neurite density index (NDI), cortical thickness (CT) and surface area (SA). Cortical regions with higher PLS1 weights (shades of yellow) have stronger genetic covariation with schizophrenia. Scatterplots show the genetic relationships between schizophrenia (y-axis, *Z*-scores from H-MAGMA analysis of schizophrenia GWAS dataset) and brain structure measured using surface area (x-axis, *Z*-scores from H-MAGMA analysis of MRI GWAS datasets) in two cortical regions, one with a low PLS1 weight (left, dark blue, *ρ* = 0.04), and one with a high PLS1 weight (right, yellow, *ρ* = 0.17). Each point represents one of 18,640 genes and higher PLS1 weighting indicates a region where the same genes were associated with both schizophrenia and cortical structure.

For SA and CT all (180) cortical areas had significantly non-zero PLS1 weights, and likewise for NDI at 179 areas, indicating that all MRI metrics were genetically covariant with schizophrenia across large areas of the cortex (Fig. 2 B). However, there was substantial regional variation of PLS1 weights for each MRI metric. The highest PLS1 weights for SA were found in insular and medial prefrontal cortex; for CT in visual, premotor and inferior parietal cortex; and for NDI in inferior frontal, inferior parietal, posterior cingulate and posterior opercular cortex (Fig. 2 B). Convergently, the anatomical patterns of genetic covariation represented by cortical maps of PLS1 weights were only weakly correlated between MRI metrics (Spearman’s *ρ*(*SA,CT*) = 0.03, *P* = 0.65; *ρ*(*SA, NDI*) = 0.17, *P*= 0.03; and *ρ*(*CT, NDI*) = 0.19, *P* = 0.01). In general, however, as illustrated in Fig. 2 B, regions with higher (positive) PLS1 weights generally showed stronger positive correlations between *Z*-scores for association with schizophrenia and *Z*-scores for association with brain MRI metrics, compared to regions with lower (negative) PLS1 weights.

We tested PLS1 weights for enrichment in relation to prior atlases of laminar differentiation [28] or functional MRI networks [29] (Methods). While the strength of genetic covariation with schizophrenia was somewhat related to cytoarchitectonically defined classes of cortical areas, we did not find any enrichment related to functional networks (Table S8-S9, Fig. S1). These results are compatible with the prior observation that genetic covariation between brain structure and schizophrenia is expressed diffusely across the cortex [6], as well as with previously published case-control MRI studies reporting widespread cortical abnormalities in schizophrenia [1].

### Genetic similarity and and structural covariance networks

We further investigated the genetic architecture of brain structure, going beyond the associations of genetic variation with each independently analysed regional MRI phenotype, to consider genetic associations with brain network phenotypes. Specifically, we analysed the relationship between inter-regional phenotypic covariance (henceforth structural covariance *SC*) estimated across N = 31,780 scans from the UK Biobank, and inter-regional genetic correlation (henceforth genetic similarity *GS*) (Methods, Fig. 3 A).

**Figure 3.**
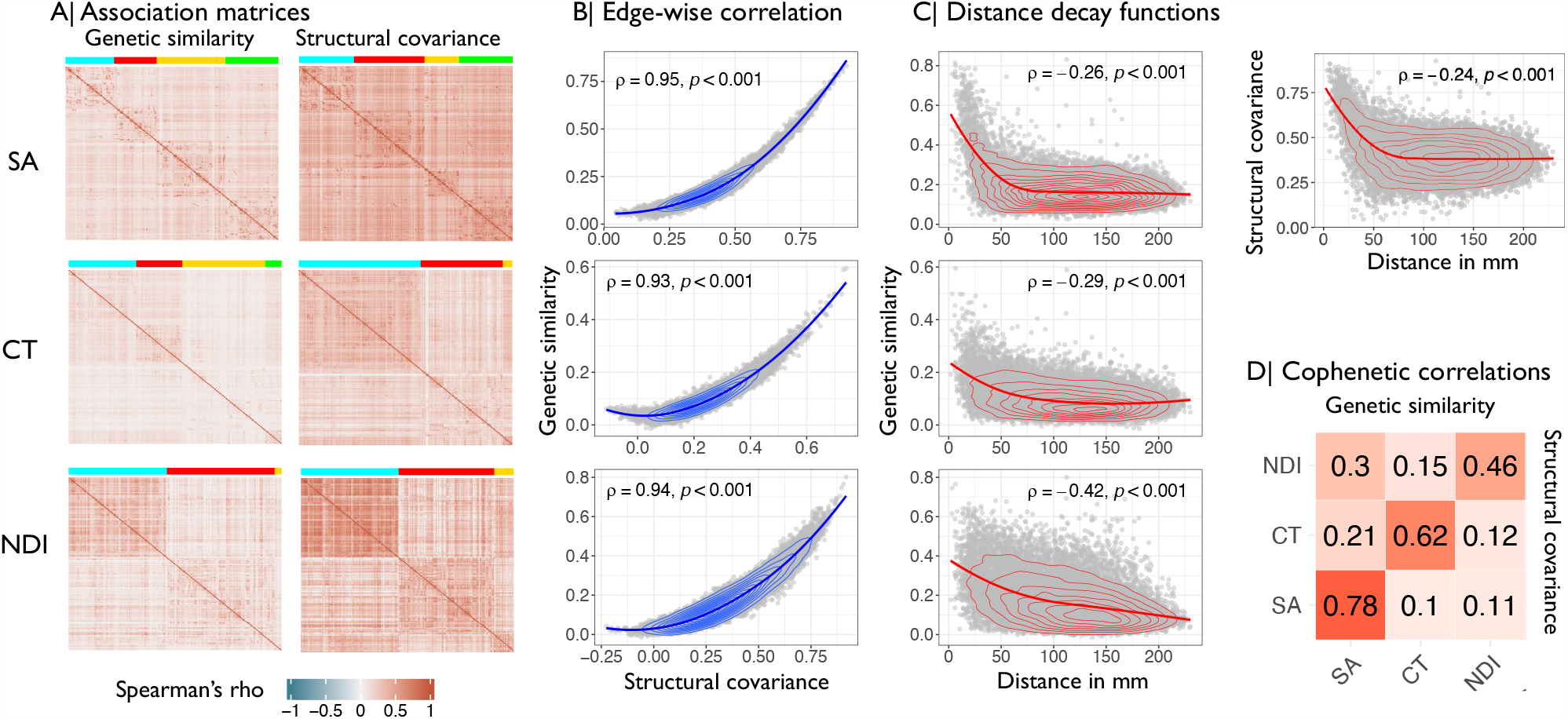
Genetic similarity and structural covariance of cortical networks. (A) Genetic similarity (left) and structural covariance (right) matrices for surface area (SA), cortical thickness (CT), and neurite density index (NDI). Brain regions are ordered according to modular decomposition based on genetic similarity and structural covariance, respectively. (B) Edge-wise correlation between genetic similarity (y-axis) and structural covariance (x-axis) matrices. (C) Correlation between genetic similarity (y-axis) and geodesic distance in millimeters (x-axis). For SA, the correlation between structural covariance and geodesic distance is also shown in the top right panel. (D) Cophenetic correlation matrix showing the similarity in hierarchical clustering of structural covariance and genetic similarity matrices. The upper triangle shows cophenetic correlations based on genetic similarity, the lower triangle is based on structural covariance, and the diagonal represents the similarity between structural covariance and genetic similarity of the same MRI metric.

In line with Cheverud’s conjecture [17], we found that there was a strong relationship between structural covariance and genetic similarity. The corresponding structural covariance (*SC*) and genetic similarity (*GS*) matrices were highly positively correlated with each other, *R*(*SC, GS*) ∼ 0.95, indicating that regions with high structural covariance had highly similar genetic profiles of association with regional variation: for SA, *R*(*SC, GS*) = 0.96; for CT *R*(*SC, GS*) = 0.93; and for NDI *R*(*SC, GS*) = 0.94 (Fig. 3 B).

Both structural covariance and genetic similarity were greatest between regional nodes separated by the shortest geodesic distances, and both declined monotonically as a function of increasing distance. For genetic similarity, the correlations with geodesic distance were: SA, *ρ* = -0.26; CT, *ρ* = -0.29; NDI, *ρ* = -0.42; all *P* < 0.0001. Whereas, for structural covariance, the correlations with geodesic distance were: SA *ρ* = -0.24; CT *ρ* = -0.3; NDI *ρ* = -0.4; all *P* < 0.0001 (Fig. 3 C, Fig. S2). Additionally, genetic similarity and structural covariance showed a consistent relationship with atlases of functional network organization [31] and cytoarchitecture [28] (Figs. S3 - S4, Table S10-S11, Methods).

Each of the *GS* and *SC* matrices was decomposed into a hierarchical cluster structure represented by a dendrogram, and the complex branching structures of the dendrograms were compared in terms of their cophenetic correlation *R*_*c*_(*GS, SC*) (Methods). For each MRI metric there was a significantly positive cophenetic correlation between the corresponding *SC* and *GS* dendrograms: for SA, *R*_*c*_(*GS, SC*) = 0.78; for CT, *R*_*c*_(*GS, SC*) = 0.62; for NDI *R*_*c*_(*GS, SC*) = 0.46; all *P* < 0.0001, by permutation tests. This level of correspondence between the detailed hierarchical structure of genetic similarity and structural covariance matrices for each metric was greater than the level of correspondence between dendrograms from pairs of *SC* matrices (*SC*_*MRI*_, *SC*_*MRI*_*′*, or pairs of *GS* matrices (*GS*_*MRI*_, *GS*_*MRI*_*′*), representing different MRI metrics: for genetic similarity, 0.15 ≥ *R*_*c*_(*GS*_*MRI*_, *GS*_*MRI*_*′*) ≤ 0.3; and for structural covariance, 0.1 ≥ *R*_*c*_(*SC*_*MRI*_, *SC*_*MRI*_*′*) ≤ 0.12, suggesting that the hierarchical clustering of structural covariance and genetic similarity networks is quite specific to each of the MRI metrics (Fig. 3 D).

We used the Louvain algorithm to resolve the modular community structure of each MRI modality and found three (for NDI) or four (for CT, SA) spatially contiguous modules of the *GS* networks, and four (SA) or three (CT, NDI) modules of the corresponding *SC* networks (Methods). The modules differed between phenotypes implying that modular structure of genetic and phenotypic brain networks is highly dependent on the underlying MRI metric (Fig. 3 A, Fig. 4 A).

**Figure 4.**
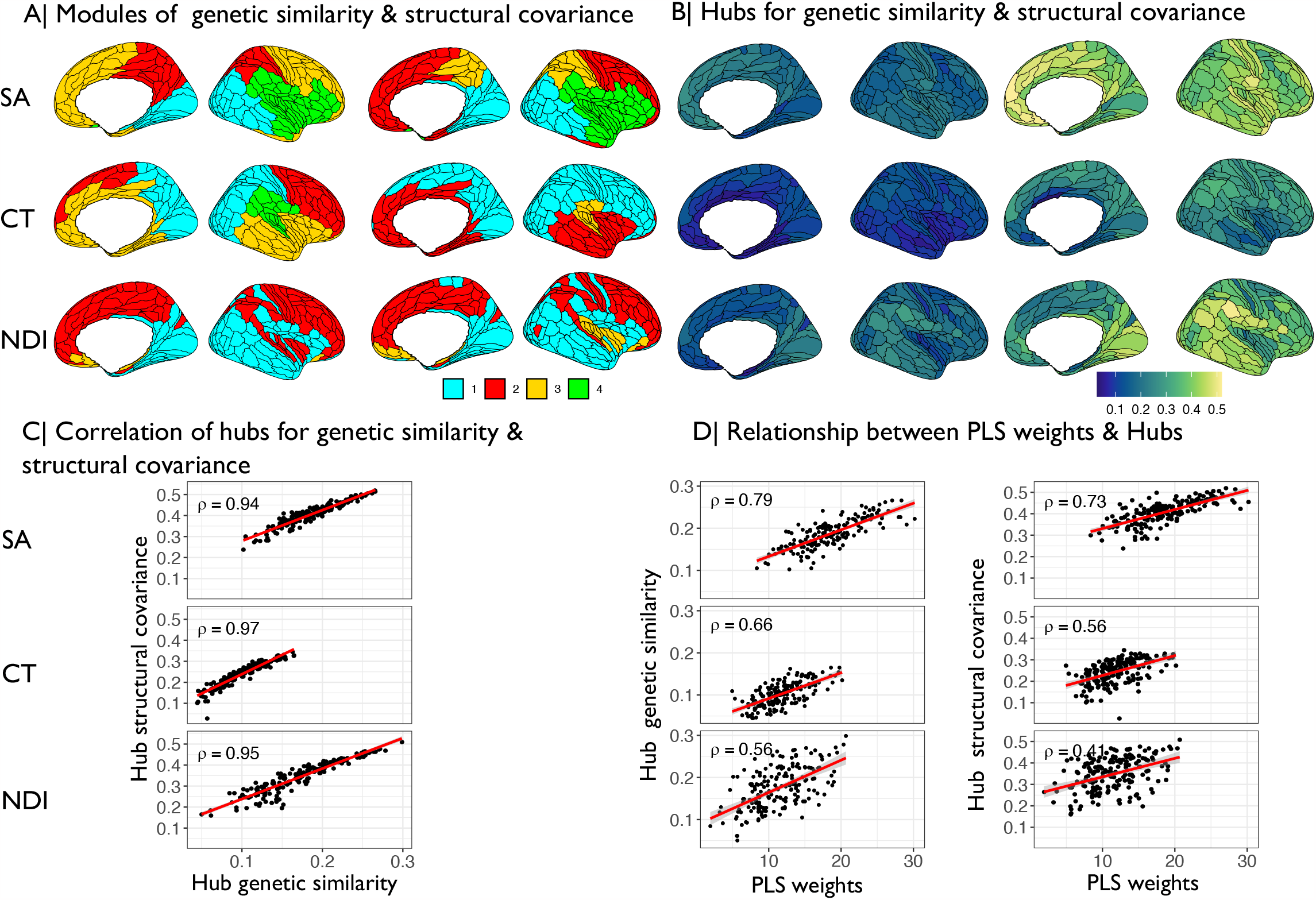
Hubs of genetic similarity and structural covariance networks are co-located and associated with pleiotropic genes. (A) Modular decomposition for surface area (SA), cortical thickness (CT) and neurite density index (NDI) based on genetic similarity matrices (left) and structural covariance matrices (right). (B) Spatial map of hub scores based on genetic similarity matrices (left) and structural covariance matrices (right). (C) Pairwise correlation between hubness of nodes in genetic similarity (x-axis) and structural covariance networks (y-axis). (D) Correlation between strength of pleiotropic gene association indexed by PLS1 weights (x-axis) and hub scores of nodes in genetic similarity networks (left) or structural covariance networks (right) (y-axis).

### Genetic relationships between schizophrenia and brain network organization

Since structural MRI covariance is normatively coupled to genetic similarity, and cortical MRI variance is associated with gene variations that are also associated with schizophrenia, we predicted that brain regions associated with schizophrenia genes might have distinctive topological profiles in the whole brain connectome represented by the structural covariance matrix. Thus, we tested if the PLS1 weights representing strength of pleiotropic association with schizophrenia-related genes at each cortical region, *PLS*1*i*, were related to the degree centrality (“hubness”) of the corresponding node in the *SC* connectome, *k*_*i*_, and/or the intra-modular and inter-modular degrees at each node.

Hubness for each brain region was defined by the weighted degree centrality, i.e., the sum of edge weights connecting each node to the rest of the network [32]. The distribution of hubness across the cortex was not strongly correlated for genetic similarity and structural covariance networks based on different MRI metrics (Fig. 4 B). However, for each MRI metric considered separately, the hub scores based on genetic similarity and structural covariance networks were very highly positively correlated (Fig. 4 C), indicating that the structural network organization closely conforms to the genetic architecture of the brain. Hub scores in structural covariance networks were positively correlated with PLS1 weights, representing the strength of pleiotropic association at each cortical area: SA *ρ* = 0.73, CT *ρ* = 0.55, NDI *ρ* = 0.4; all *P* < 0.0001 (Fig. 3 D). We additionally computed intra-modular and inter-modular degree for each brain region based on structural covariance matrices (Methods). PLS1 weights were positively correlated with intra- and inter-modular degree for all MRI metrics (intra-modular degree SA *ρ* = 0.55, CT *ρ* = 0.48, NDI *ρ* = 0.22; inter-modular degree SA *ρ* = 0.39, CT *ρ* = 0.38, NDI *ρ* = 0.73; all *P* < 0.0001), indicating that stronger pleiotropic gene associations are found in in regions that are relevant for local connectivity as well as long-range connections between modules (Fig. S5 A-B)

### Pleiotropic genes mediating covariation between schizophrenia and regional MRI metrics

To identify genes which made the greatest contribution to whole genome covariation between schizophrenia and regional brain MRI metrics, we focused on the *T* and *U* scores derived from PLS analysis (Methods and Fig 2) and the correlation between them, *R*(*T,U*).

The strength of pleiotropic association with schizophrenia, across all 18,640 genes, was greatest for SA (*R*(*T,U*) = 0.24), then for CT (*R*(*T,U*) = 0.23), and then NDI (*R*(*T,U*) = 0.17); see Figure 5 A. To assess the influence of an individual gene on these whole genome relationships, we used a leave-one-out (LOO) strategy and computed Δ(*R*(*T,U*)), i.e. the difference between the original and the LOO *R*(*T,U*), for each gene. Genes that make the greatest individual contribution to pleiotropic association will have the largest positive values of Δ(*R*(*T,U*)) (Methods).

**Figure 5.**
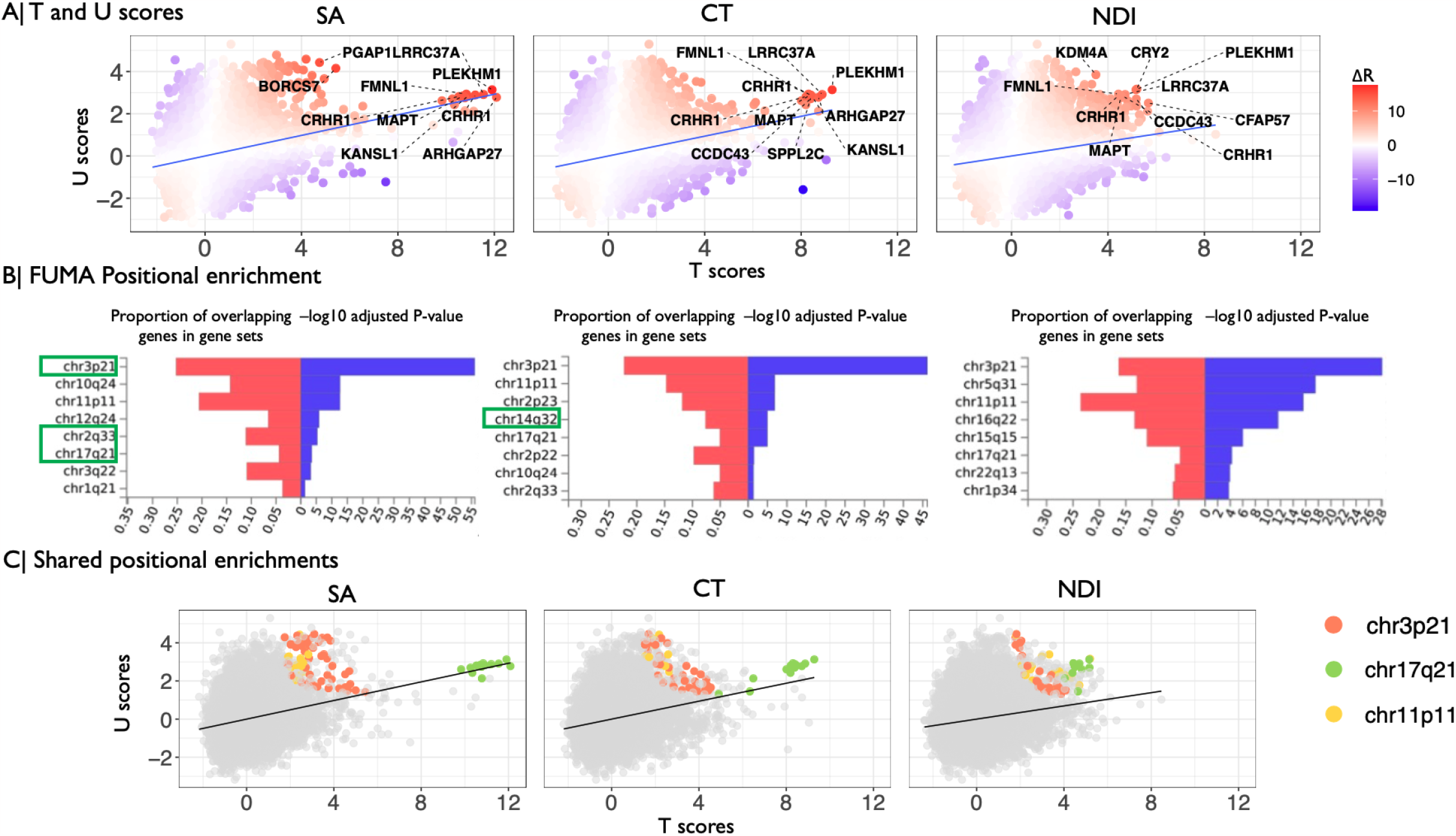
Genes contributing to genetic covariation between schizophrenia and regional MRI metrics. (A) Scatterplot of *T* scores (*x*-axis) versus *U* scores (*y*-axis) for each of 18,640 protein-coding genes, derived from their weights on the first PLS component. The *T* score is the weight of each gene on the MRI metric; the *U* score is the weight of each gene on the association with schizophrenia; and the correlation between *T* and *U* scores, *R*(*T,U*), quantifies the strength of genetic relationship between brain structural and schizophrenia phenotypes. Each gene is colour-coded according to its value of Δ(*R*(*T,U*)) which indicates the positive (red) or negative (blue) magnitude of its influence on the whole genome relationship between schizophrenia and each MRI metric. The top ten genes with the largest positive leave-one-out scores for Δ(*R*(*T,U*)) are annotated, including *PLEKHM1, FMNL1, LRRC37A, MAPT, KANSL* and *CRHR1*, all located within the 17q21.31 region. We note that these genes were also identified by the intersection analysis of genes significantly associated with both MRI metrics and schizophrenia. (B) Significant positional enrichment of 185 genes (top 1%) with highest Δ(*R*(*T,U*)) scores based on FUMA. For example, the genes most strongly contributing to genetic covariation between SA and schizophrenia were positionally enriched at chromosome 2q33, whereas genes contributing to covariation between CT and schizophrenia were enriched at chromosome 14q32. The red bars show the proportion of co-located genes according to the size of each gene-set; the blue bars indicate -log10 *P*-values adjusted for the number of tested gene-sets. Chromosomal locations showing significant local genetic correlations based on LAVA are highlighted in green boxes. (C) Scatterplot of *T* scores versus *U* scores, exactly as shown in (A) except that genes are colour-coded according to their location in three genomic regions that were positionally enriched for all MRI metrics.

We functionally characterised the top 1% (or 3%) of genes with largest positive Δ(*R*(*T,U*)) for each MRI metric (Table S14). These sets of 185 genes were significantly enriched for constrained genes, which are intolerant of damaging variants and have previously been associated with brain structure [3] and schizophrenia [33]. The pleiotropic gene sets were not strongly enriched for cell type-specific genes (Fig. S7, Table S15) but we identified 33 significant GO enrichments (six for SA, 22 for CT and five for NDI) (Table S16) related to neurodevelopmental processes including neurogenesis (SA GO:0022008, *P* = 0.001), nervous system development (CT GO:0007399, *P* = 0.002), glial cell development (CT GO:0048468, *P* = 0.009) and neuron projection development (NDI GO:0010975, *P* = 0.03).

We tested whether pleiotropic genes were over-represented in discrete genomic regions using hypergeometric testing implemented in FUMA [34] (Methods). We identified eight loci of significant positional enrichment for one or more of the 3 MRI metrics, most of which were specific to a single metric (Fig.5 B). However, genes pleiotropically associated with all 3 MRI metrics were positionally enriched at the same three genomic regions: chromosome 3p21 (chr3:43,700,001-54,400,000), chromosome 17q21 (chr17:38,100,001-50,200,000) and chromosome 11p11 (chr11:43,500,001-53,700,001). The strongest positional enrichment for each MRI metric was on chromosome 3p21, suggesting that genes concentrated at this location strongly influence the genetic relationship between schizophrenia and all three brain MRI metrics (Fig.5 C).

Since the loci of positional enrichment identified using FUMA were relatively long (≥ 5Mb ≤ 25 Mb), we also estimated local genetic correlations at these positions to identify shorter genomic segments (∼ 1Mb) mediating genetic covariation between MRI metrics and schizophrenia using LAVA (Local Analysis of [co]Variant Association) [35] (Methods). Within four of the positionally enriched loci identified by FUMA, we were able to further narrow down the signal to smaller subregions (≥ 1Mb ≤ 2 Mb), e.g., chromosomal loci 14q32.2 - 14q32.31 and 14q32.13 for CT, and 3p21.2 - 3p21.1, 2q33.1 and 17q21.31 for SA (Fig.5 B). We did not find any significant local genetic correlations for NDI (Table S16).

## Discussion

We have identified genes significantly associated with one or more of three relatively distinct regional MRI metrics (surface area, cortical thickness and neurite density index) and reported widespread genetic covariation between schizophrenia and these measures of cortical structure. We have shown that structural covariance networks are closely coupled to genetic similarity networks for each metric and that genetic associations with both schizophrenia and brain structure were strongest in cortical areas constituting hubs of the whole brain connectomes. Finally, we have shown that the genes making the greatest individual contributions to whole genome covariation between schizophrenia and MRI metrics were enriched for mutation-intolerant genes, functionally specialised for neurodevelopmental processes, and positionally enriched in three plausible genomic regions relevant for schizophrenia.

### Genetic associations with MRI metrics

Most genes associated with brain structure had specific effects on one of the three MRI metrics, supporting the premise that SA, CT and NDI are relatively independent cortical phenotypes that might be influenced by distinct genetic mechanisms [2, 3]. However, genes associated with each MRI metric broadly shared transcriptional trajectories that peaked during mid-late periods of fetal life, which is well-recognised as a key neurodevelopmental period for the formation of upper layer neurons and neuronal differentiation (including axonogenesis and dendritic arborization) [36]. Several genes were generically associated with all three MRI metrics, including 16 genes located within the 17q21.31 region, a known inversion polymorphism with a complex LD structure [25], which has previously been associated with diverse imaging phenotypes including left-right asymmetry [37], white matter microstructure [38], intracranial volume [39], and functional connectivity [40]. Genes associated with SA and CT were also enriched for biological processes including microtubule function, which is known to be important for neurogenesis, neuronal migration and axon generation [41]. Collectively these results indicate that both unique and shared genetic associations with brain structural phenotypes likely represent the effects of genetically controlled programmes of gestational and early post-natal development of the cortex.

### Genetic covariation between schizophrenia and regional MRI metrics

The prevailing hypothesis for the pathogenesis of schizophrenia is the neurodevelopmental model, positing that genetic and environmental factors perturb normal processes of brain development, including formation of synaptic connections and axonal projections between cortical areas, with anatomically distributed effects on adult brain connectivity [8, 42, 43]. In support of this model, we first demonstrated that there was greater intersection than expected by chance between genes associated with brain structure and schizophrenia. For example, about 9% of the 586 genes significantly associated with schizophrenia were also significantly associated with normative variation in surface area of one or more cortical regions. We subsequently used partial least squares to investigate genetic covariation over the whole genome and to comprehensively map cortical locations of significant genetic covariation with schizophrenia. We found evidence for significant pleiotropic association with both schizophrenia and surface area or cortical thickness of all cortical areas, and for NDI at more than half (179/180) of all cortical areas, indicating that genetic risks for schizophrenia are also associated with widespread variations in local cortical anatomy. This observation is consistent with global or extensive regional differences in cortical structure reported by prior case-control MRI studies of schizophrenia [1]. However, the cortical patterns of genetic covariation with schizophrenia were only weakly correlated between MRI metrics and there was considerable inter-areal variation in the strength of pleiotropic association for each metric. For example, genetic covariation with surface area was greatest in paralimbic areas of cortex that have previously been associated with polygenic risk for schizophrenia [6]. In short, we found convergent evidence from two technically distinct analyses for genetic covariation between schizophrenia and cortical structure, although the strength of covariation varied between cortical areas and MRI metrics.

### Genetic and phenotypic brain network concordance

We used between-subject covariance of regional MRI metrics as a proxy for anatomical connectivity between each pair of cortical areas in a structural covariance network constructed from all the scans of each MRI metric [13]. Each of these three connectomes could then be compared to the corresponding genetic similarity matrix representing the pair-wise correlations between regional metrics in terms of their whole genome profiles of association with each of 18,640 genes. For each MRI metric, there was a remarkably strong degree of concordance between structural covariance and genetic similarity networks. Structural covariance and genetic similarity were highly correlated over all edges in both networks (or all elements in both association matrices) and were both strongest between anatomically neighbouring regions with monotonically decreasing strength as a function of increasing geodesic distance between regional nodes. The neurobiological substrate of structural covariance networks has long been debated [13] but these results strongly suggest that structural covariance between regions represents close equivalence in the genetic determinants of their development. For example, positive covariance in thickness or surface area between two association cortical areas in a sample of adult brain scans likely reflects that those two areas have followed very similar, genetically-controlled trajectories of development since mid-fetal life.

Genetic similarity and structural covariance networks for each metric were also highly concordant in terms of some of their topological properties. For example, the hubs of the cortical thickness covariance network, which are often interpreted as cortical areas with a high density of axonal connectivity to other areal nodes in the connectome [13], were generally also hubs in the genetic similarity network derived from GWAS data on cortical thickness at each node. Hubs in the genetic similarity matrix are cortical areas that share a whole genome profile of genetic association in common with many other areas of cortex, and are therefore putatively under the same genetic controls throughout development. So one interpretation of the strong coupling between genetic and structural covariance network hubs is that cortical areas are more likely to be axonally connected if their differentiation and development is controlled by similar genetic mechanisms. There is convergent evidence in support of this “grow together, wire together” model from recent studies in animals and humans demonstrating that the probability of inter-areal axonal projections is increased between cortical areas with similar cytoarchitectonics [44] or whole genome transcription profiles [45].

In this context, it is notable that regions with the highest genetic covariation with schizophrenia genes were hub regions of the corresponding connectomes. One interpretation is that the neurodevelopmentally-enriched genes associated with schizophrenia also have an important normative role in development of anatomical inter-connectivity between cortical areas, and are therefore most strongly associated with the most highly connected hub nodes of the connectome. The flip side of this interpretation is that genetic variants associated with schizophrenia may cause atypical development of brain network hubs, in particular, with emergent consequences for “higher order” cognitive processes that are often impaired in schizophrenia [46]. Consistent with this interpretation, previous studies have shown that whole-genome transcriptional profiles co-located with adolescent myelination of structural covariance network hubs were enriched for genes associated with schizophrenia [47]; and that schizophrenia cases, compared to healthy controls, had reduced degree of cortical hubs in morphometric similarity networks that were co-located with cortically patterned expression of schizophrenia-related genes [16].

### Chromosomal clusters of genes pleiotropically associated with schizophrenia and brain structure

Across all MRI metrics we found that the gene sets with the strongest influence on genetic covariation were enriched for constrained genes that are intolerant of damaging variants. This finding aligns with previous evidence that the genetic architecture of schizophrenia is substantially driven by constrained genes [20, 48]. We also showed that pleiotropic genes were plausibly enriched for neurodevelopmental and glial ontology terms, echoing previous work highlighting the role of glial cells in typical and atypical development of brain networks [49, 50].

However, one of the most striking, robust and contextually plausible characteristics of the genes associated with both brain structure and schizophrenia was their concentration or clustering on a few chromosomal regions, in particular, chromosome 3p21, chromosome 11p11 and chromosome 17q21. Genetic variation in the 17q21 region has been associated with various measures of brain structure [37, 38, 39, 40] as well as schizophrenia [20, 51] and other psychiatric (e.g. autism spectrum disorder [52]) and neurodegenerative disorders (e.g. Alzheimer’s disease [53]). Using LAVA we were able to further narrow down the signal for surface area to a subregion of 17q21.31 spanning ∼ 1.4 Mb. This locus harbors a inversion polymorphism and includes *PLEKHM1, MAPT, KANSL1* and *CRHR1. CRHR1* encodes the main receptor of corticotrophin-releasing hormone and has recently been highlighted in a study of shared genetic effects on schizophrenia and subcortical volumes [54]. *MAPT* encodes microtubule-associated protein tau, which is known for its role in axonal transport and neurite outgrowth and has previously been associated with schizophrenia and structural MRI metrics [7]. *PLEKHM1* is involved in autophagy [55], a process that has been suggested to have a key role in the pathophysiology of schizophrenia [56]. Chromosome 11p11 also harbours genes that have been previously associated with schizophrenia and/or brain structural phenotypes, including *CHRM4, MDK, AMBRA1* and *HARBI1* [20, 57]. For example, *AMBRA1* has a major role in the development of the nervous system during early fetal life and has been implicated in neuronal migration, differentiation and maturation [58]. *CHRM4*, encoding the muscarinic acetylcholine receptor M4, has been linked to the genetic risk for schizophrenia, with reduced hippocampal expression in post mortem cases [59], and positive clinical trials of M4 agonists for the treatment of schizophrenia [60, 61]. Finally, chromosome 3p21 has also previously been associated with schizophrenia risk [20, 33, 62], other psychiatric disorders [63], cognition [64] and measures of brain structure [40]. Using LAVA we identified the subregion 3p21.2 - 3p21.1, spanning ∼ 2.1 Mb. This region harbors genes including *ITIH4, NEK4, GNL3* and *PBRM1. ITIH4* encodes inter-alpha-trypsin inhibitor heavy chain 4 and was previously found to be associated with schizophrenia risk and intracranial volume and is also implicated in inflammatory responses [5]. Over-expression of either *NEK4* or *GNL3*, or knockdown of *PBRM1* (encoding serine/threonine protein kinase, guanine nucleotide-binding protein-like 3, or polybromo-1, respectively) was shown to reduce mushroom dendritic spine density in cortical neurons in rats [62], mirroring the dysmorphic dendritic arborization observed in schizophrenia [65].

### Strengths and limitations

Using diverse methodological approaches, we were able to identify three genomic regions that plausibly influence schizophrenia and regional brain structure. While some of the identified genes are well characterised (e.g. *MAPT, CRHR1*), the function of other identified genes (e.g. *KANSL1, PLEKHM1* and *LRRC37A*) are not fully understood. Future experimental studies investigating the biological function of the identified genes are required to understand the complex, biological relationship between brain structure and schizophrenia.

### Conclusion

Common variants in protein-coding genes pleiotropically associated with both schizophrenia and regional brain structure, especially in hub nodes of the cortical connectome, were functionally specialised for neurodevelopmental processes and positionally concentrated on three plausible genomic regions. These results support a pathogenic model of schizophrenia as the clinical expression of a genetically driven disorder of brain network development that originates in fetal and early post-natal life.

## Methods and Materials

### GWAS

We accessed recently published GWAS summary statistics on three imaging phenotypes including surface area, cortical thickness and neurite density index, measured at 180 brain regions (3 × 180 GWAS). The GWASs were based on 36,843 subjects from the UK Biobank [66] and the Adolescent Brain and Cognitive Development (ABCD) study [67]. Details can be found in [3]. To investigate genetic overlaps with schizophrenia, we accessed GWAS summary statistics based on 69,369 schizophrenia cases and 236,642 controls [20].

### H-MAGMA

Single-nucleotide polymorphisms were mapped to genes using the default settings in H-MAGMA with fetal and adult brain Hi-C annotation datafiles provided by the developers of H-MAGMA (https://github.com/thewonlab/H-MAGMA) and the reference data file for a European ancestry population downloaded from https://ctg.cncr.nl/software/magma. Since the genetic signal for both schizophrenia and brain structure are enriched for regulatory regions active in the fetal brain [3, 68], we report findings based on fetal Hi-C datasets. We restricted our analyses to protein-coding genes and excluded genes within the major histocompatibility region due to the complexity of linkage disequilibrium, which can override the overall pattern [21].

### Genetic effects on regional MRI metrics

#### Identification of genes associated with MRI metrics

To investigate the genetic profiles of regional brain structure we identified a set of significant genes for each MRI metric. To account for multiple comparison correction, we performed matrix decomposition within each MRI metric to identify the number of independent phenotypes (*N*_*phenotypes*_)[69]. Modality-wide significance thresholds were then calculated for each MRI metric by using Bonferroni correction of the total number of tests (*P* = 0.05 / 18,640 *genes* x *N*_*phenotypes*_) [3]. We identified 78 independent phenotypes for SA, 113 for CT and 74 for NDI. Thus, the modality-wide significance thresholds varied between imaging modalities (SA *P* ≤ 3.44 × 10^−8^; CT *P* ≤ 2.37 × 10^−8^; NDI *P* ≤ 3.62 × 10^−8^).

#### Gene Ontology and Developmental expression profiles

We used the R package gProfiler2 for conducting GO enrichment analysis which resembles gene set enrichment analysis, and focused enrichment tests on biological processes [70]. We investigated developmental expression profiles of genes that were significantly associated with regional brain structure. To ensure that developmental expression trajectories were not biased by the developmental stage from which Hi-C data were obtained, we created lists of significantly associated genes that were either identified using fetal or adult Hi-C data [21]. A spatiotemporal transcriptomic atlas from PsychEncode [26] was used to obtain cortical expression profiles across nine developmental stages (window 1: PCW 5-9, window 2: PCW 12-13, window 3: PCW 16-18, windows 4: PCW 19-22, window 5: PCW 35-PY 0.3, window 6: PY 0.5-2.6, window 7: PY 2.8-10.7, window 8: PY 13-19, window 9: PY 21-64). This data set contains expression values from multiple brain regions and we selected transcriptomic profiles of the cerebral cortex in line with our imaging analyses, which were conducted in cortical regions only. Expression values were log-transformed and centered to the mean expression level for each sample using `scale(center = T, scale= F)+1` function in R. We measured the average expression values of the entire gene set [71]. To do this, genes associated with regional brain structure were selected for each sample and their average centered expression values were calculated and plotted. Additionally, we investigated the effect of developmental window (prenatal expression included windows 1-4, postnatal expression included window 6-9) on gene expression using linear mixed effect models, with a fixed effect of developmental window and gene length, and a random effect of brain region. Results were FDR corrected across all MRI metrics.

### Genes associated with schizophrenia

To identify genes that were significantly associated with schizophrenia we mapped GWAS summary statistics based on 69,369 schizophrenia cases and 236,642 controls [20] to genes using H-MAGMA. We restricted genes to protein coding genes and excluded genes located in the major histocompatibility complex (MHC) region. We performed Bonferroni correction for all 18,610 genes leading to a significance threshold of *P* ≤ 2.69 × 10^−6^. We used hypergeometric testing implemented in the R package GeneOverlap [72] to test for significant overlap between schizophrenia-associated genes and MRI metric-associated genes and performed permutation testing (10,000 permutations) to test whether this overlap is non-random. To investigate enrichments for GO terms and developmental trajectories we used the same pipeline as outlined above.

### Partial least squares regression: PLS weights

For each MRI metric, we extracted the first PLS (PLS1) component using the `plsdepot` package in R [73]. The significance of PLS1 was estimated by comparing the empirical variance explained by each component to a null distribution, i.e. the distribution of variance explained when permuting Y 1000 times. Additionally, we tested whether PLS weights were spatially enriched in known cortical atlases of network organisation (Yeo networks [29]) or cytoarchtectonic classes (mesulam classes [74]), and in identified modules by spin permutations. Spatial permutation testing (spin-tests) were used to control for spatial autocorrelation in PLS maps [75, 76]. Results were FDR-corrected for each MRI metric and cortical atlas.

### Structural covariance and genetic similarity networks

To generate structural covariance matrices, we accessed imaging data from the UK Biobank and focused on a subset of N = 40,680 participants for each of whom complete genotype and multimodal MRI data were available for download (February 2020) [66]. Details on processing pipeline and exclusion criteria and are described in SI Methods. After quality controls N = 31,780 MRI scans were available for each MRI metric. All samples compromised ∼ 53% female, ∼ 47% male participants aged 40-70 years, with mean age 55 years (SD = 7.4 years). Thus, structural covariance matrices were based on a subset of subjects that were part of the MRI GWAS [3]. For each MRI metric, we estimated the pair-wise correlation of regional phenotypes for all pairs of 180 regions over all N = 31,780 MRI scans from the UK Biobank, to constitute a symmetric, signed { 180 *×* 180 } structural covariance matrix. Each unique, off-diagonal element of this matrix, *SC*_*i, j*_ can be regarded from the perspective of graph theory as the weight of an undirected edge between regional nodes *i* and *j* in a whole brain structural connectome. We also estimated the similarity of genetic association between each possible pair of regional nodes *i* and *j* in terms of Spearman’s correlation, *ρ*_*i, j*_, between their gene-level association statistics (*Z*-scores derived from H-MAGMA) to constitute a symmetric, signed { 180 *×* 180 } genetic similarity (*GS*) matrix.

#### Geodesic distance, functional networks and cytoarchitectonic classes

We investigated the association between genetic similarity (or structural covariance) and geodesic distance, known functional networks and cytoarchitectonic classes. Geodesic distance was defined as the length of the shortest path between each pair of regions along the cortex measured in millimeters and correlated (Spearman’s correlation) with SC and GS respectively. We tested whether genetic similarity and structural covariance were higher within resting-state functional networks [29] or cytoarchitectonic classes [74] by comparing the average genetic similarity (resp. structural covariance) of regions within networks (resp. classes) to regions between networks (resp. classes). We used spin-tests (1000 permutations) to test the significance of the co-location of two cortical maps while accounting for spatial-correlation between brain regions [75, 76]. To control for multiple testing, we performed FDR-correction across all networks (resp. classes).

#### Cophenetic correlations

Cophenetic correlation coefficients were used to quantify the similarity in network structure based on GS and SC within and between phenotypes. To this end, Euclidian distance matrices were calculated using the `dist()` function, and `hclust(method = “ward)` was used to generate dendrograms for each phenotype. Cophenetic correlation coefficients were computed using the `cor_cophenetic()` function from the dendextend R package [77]. To test for statistical significance, we generated a null distribution of cophenetic correlation coefficients by permuting over the labels of one dendrogram 1000 times while keeping the dendrogram topologies constant.

#### Modular decomposition, intra- and inter-modular degree

To characterise the network topology of structural covariance and genetic similarity matrices, we created weighted, undirected network graphs based using `igraph` in R [78]. Modules were identified using the `cluster_louvain()` function, which implements a multi-level modularity optimization algorithm for finding community structure [79]. For each node, intra-modular degree was computed as the mean weighted degree over all nodes in the same module. Inter-modular degree was estimated as the mean weighted degree over all nodes outside the module [32].

### Partial least squares regression: T and U scores

To identify which genes most strongly impact the relationship between regional brain structure and schizophrenia, we investigated *T* scores and *U* scores, quantifying the contribution of each gene to the covariance between brain structure and schizophrenia. More formally, *T* is defined as the product of *Xw*_*X*_ and *U* the product of *Yw*_*Y*_. PLS maximises the covariance of *X* and *Y* with:

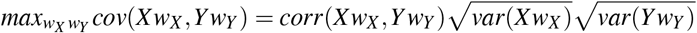

Thus, covariance is a product of three terms: correlation (between *T* and *U* scores) and two standard deviations (of *T* and *U* scores). The correlation term informs about the true association between *T* and *U* (the standard deviations reflect variances within *X* and *Y*), and given that correlation does not have a directionality (i.e., *T* and *U* are equally important), both *T* and *U* scores should be considered to select genes [27].

We used a leave-one-out (LOO) strategy, systematically re-estimating *R*(*T,U*) after the exclusion of each gene in turn from the *X* and *Y* variables used for PLS estimation of *T* and *U* scores. This procedure resulted in 18,640 LOO estimates of the genetic relationship between brain MRI and schizophrenia phenotypes, each of them estimated after leaving out a single gene, *R*_*LOO*_(*T,U*). The influence of each gene on the whole genome relationship was then defined by Δ(*R*(*T,U*)) = *R*(*T,U*) −*R*_*LOO*_(*T,U*), i.e., genes that make the greatest individual contribution to the genetic relationship will have the largest positive values of Δ(*R*(*T,U*)) [27].

#### Constrained genes, cell-type enrichment, gene ontology and positional enrichment

To characterise genes that influence the covariation between schizophrenia and brain structure we investigated enrichments for constrained genes, cell types, biological processes and chromosomal positions. All enrichment anlayses were performed on the top 1% of genes with the highest Δ(*R*(*T,U*)) values and repeated on the top 3% of genes with the highest Δ(*R*(*T,U*)) values. We investigated enrichment in mutation-intolerant genes (pLOEUEF < 0.37) identified by Karczewski et al. [48] using logistic regression after accounting for log transformed gene length as a covariate. Since MRI-associated genes were highly expressed during mid-gestation, we used single cell RNA sequencing data from the mid-gestation period [80]. Specifically, expression values per cell were log-transformed and normalised. Mean cell type specific expression was divided by the average expression of genes in all cell types to calculate relative cell type expression. The average centred expression values of genes associated with each MRI modality were calculated for each cell type, and we performed linear regression analyses controlled for log transformed gene length to assess significance [3, 21]. Results were FDR corrected across cell types. We used the R package gProfiler2 for conducting GO enrichment analysis and focused on biological processes [70]. Positional enrichments were performed using FUMA (funtional mapping and annotation of genome-wide association studies). FUMA applies hypergeometric tests to investigate whether genes of interest are overrepresented in predefined chromosomal positions (MSigDB C1) while accounting for multiple comparison correction [34].

#### Local genetic correlations

We used LAVA (Local Analysis of [co]Variant Annotation) to estimate the local SNP heritability and the genetic correlation between the schizophrenia GWAS and GWAS results of brain regions with the highest PLS weights (i.e the region showing the highest genetic covariation with schizophrenia). To account for potential sample overlap we estimated the intercepts from bivariate LD Score Regression as suggested by LAVA. Since the GWAS summary statistics were from European samples, we used the European panel of phase 3 of 1000 Genome as a LD reference. LAVA splits the genome into 2,495 non-overlapping and broadly LD independent loci [35]. Since our primary goal was to identify sub-regions within the enriched FUMA positions, we restricted our analysis to loci that lied within the enriched positions leading to 71, 66 and 54 loci of interest for SA, CT, NDI. For each phenotype pair, genetic correlation analysis was only performed for loci in which both phenotypes exhibited significant SNP heritability signal at P < 0.05/71 for SA, P < 0.05/66 for CT, P < 0.05/54 for NDI, resulting in 20, 14 and six tests for SA, CT and NDI respectively. To identify significant genetic correlations we used a Bonferroni-corrected P value threshold of *P* < 0.05/20 for SA, *P* < 0.05/14 for CT, and *P* < 0.05/6 for NDI.

## Supporting information

Supplementary Information and Results

## Data Availability

All data produced in the present study are available upon reasonable request to the authors.

## Acknowledgements

E.-M.S. is supported by a PhD studentship awarded by the Friends of Peterhouse. This research was co-funded by the National Institute of Health Research (NIHR) Cambridge Biomedical Research Centre and a Marmaduke Sheild grant to R.A.I.B. and V.W. E.T.B. is an NIHR Senior Investigator. R.A.I.B. is supported by a British Academy Post-Doctoral fellowship and the Autism Research Trust. The views expressed are those of the author(s) and not necessarily those of the NHS, the NIHR or the Department of Health and Social Care. This research was possible due to an application to the UK Biobank (project 20904). We thank Dr Agoston Mihalik for his advice on partial least square regression analysis.

## Author contributions statement

E-MS, RAIB, VW, and ETB designed research. LD advised on PLS analysis. HW advised on research design. E-MS analyzed data. E-MS and ETB wrote the paper.

## Data availability

Summary statistics on cortical MRI phenotypes are available for download on https://portal.ide-cam.org.uk/overview/483. Summary statistics for schizophrenia can be accessed from the Psychiatric Genomics Consortium https://pgc.unc.edu/for-researchers/download-results/. Imaging data may be requested through the UK Biobank database https://www.ukbiobank.ac.uk/. Spatiotemporal gene expression data can be accessed from PsychENCODE http://development.psychencode.org/. Cell type specific expression data can be downloaded from http://solo.bmap.ucla.edu/shiny/webapp/. Information on constraint genes can be accessed from gno-mAD https://gnomad.broadinstitute.org/downloads#v2-constraint.

## Code availability

The code will be made publicly available upon final acceptance of the manuscript.

