## Supplementary Information and Results for "The genetic relationships between brain structure and schizophrenia"

### 1 SI Methods

#### 1.1 Imaging acquisition and preprocessing

To generate structural covariance matrices we accessed imaging data from the UK Biobank [1]. We focused on a subset of N = 40,680 participants for each of whom complete genotype and multimodal MRI data were available for download (February 2020). MRI data acquisition has been described in detail elsewhere [2]. In brief, MRI data was collected on a 3T Siemens Skyra scanner (Siemens, Munich, Germany) using a 32-channel receive head coil. T1-weighted images were acquired using a 3D MPRAGE sequence with the following key parameters; voxel size 1x1x1mm, TI/TR = 880/2000 ms, Field-of-view = 208x256x256 matrix, scanning duration: five minutes. The diffusion imaging data was acquired using a monopolar Steejskal-Tanner pulse sequence and multi-shell acquisition (b=0s/mm2, b=1,000s/mm2, b=2,000s/mm2) with the following key parameters; voxel size 2x2x2mm, TE/TR = 92/3600 ms, Field-of-view = 104x104x72 matrix and scanning duration = seven minutes [2].

We followed preprocessing steps outlined in [3]. In brief, minimally processed T1- and T2-FLAIR- weighted MRI data (and DWI data) were downloaded from UK Biobank (application 20904) and further processed with Freesurfer (v6.0.1) [4] using the T2-FLAIR weighted images to improve pial surface reconstruction. Preprocessing steps included bias field correction, registration to stereotaxic space, intensity normalization, skull-stripping, and grey/white matter segmentation; Following reconstruction, the Human Connectome Project (HCP) parcellation [5] was aligned to each individual image and regional metrics were estimated for 180 bilateral cortical areas. Neurite orientation dispersion and density imaging (NODDI) reconstruction was performed using the AMICO pipeline [6].

We excluded participants with incomplete MRI data and we additionally excluded participants who were robustly defined as outliers by global or regional metrics more than 5 times the median absolute deviation from the sample median ($\pm$ 5 MAD). For CT and SA this lead to 31,780 subjects and 31,797 subjects for NDI.

### 2 SI Results

#### 2.1 Enrichment of PLS weights in known cortical atlases

As outlined in the main text we investigated whether PLS weights were related to two predefined and one data driven cortical atlases. Specifically, we tested for enrichment in Mesulam classes [7], Yeo networks [8] and modules based on structural covariance and genetic similarity networks. Distributions were generated by spin-permutation. PLS1 weights were higher than expected by chance in the paralimbic class for SA ($Z = 2.77$, $P < 0.05$); in the heteromodal class ($Z = 2.15$, $P < 0.05$) and idiotypic class of cortex ($Z = 2.61$, $P < 0.05$) for CT; and in the heteromodal class for NDI ($Z = 2.69$, $P < 0.05$). We did not find any significant enrichments of PLS1 weights for functional networks. However, PLS weights were significantly enriched for CT in module two (Fig. S1).

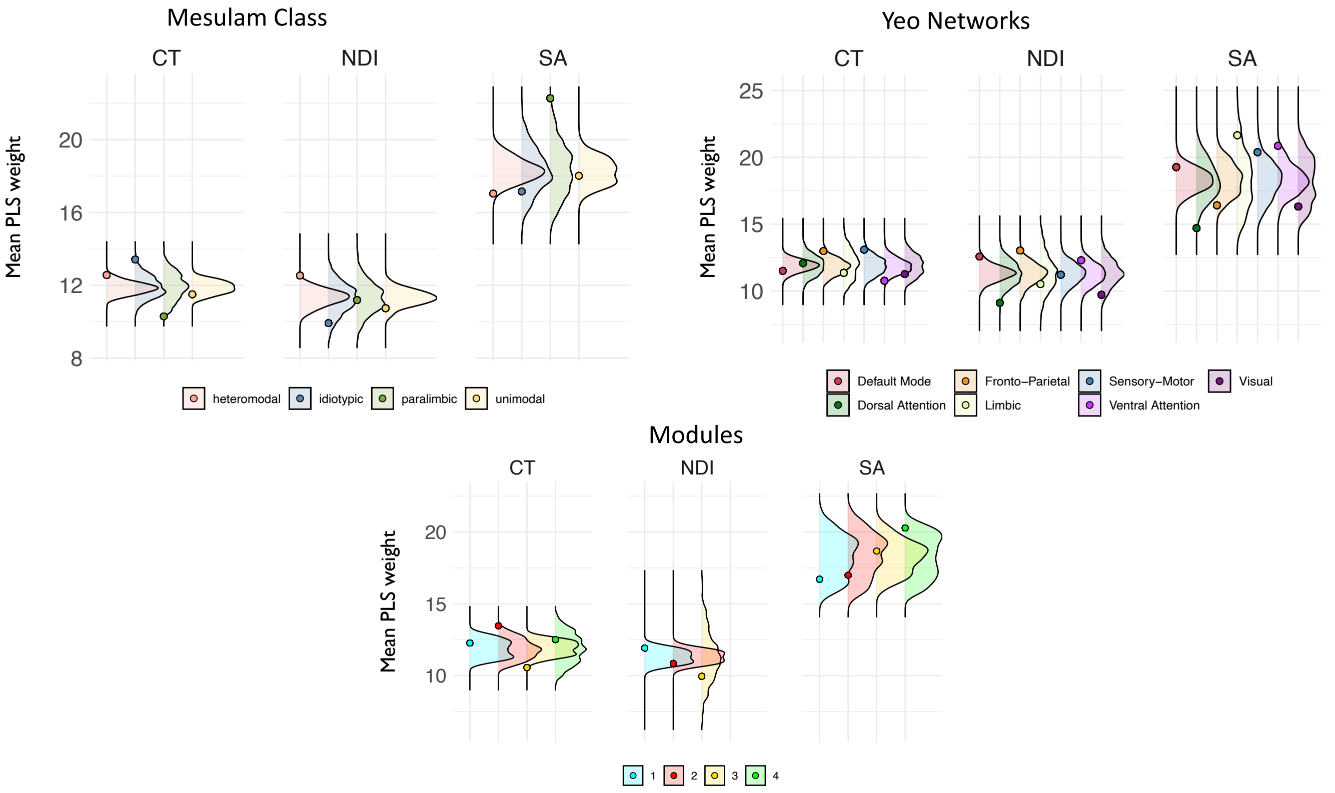

**Figure S1.** Enrichment of PLS weights for cortical thickness, neurite density index and surface area in mesulam classes, yeo networks and genetically informed modules. Distributions of mean PLS weights were generated using spin-permutations. Circles indicate the true value.

#### 2.2 Normative structural covariance and distance

Similar to genetic similarity, structural covariance was negatively associated with geodesic distance. Shown are distance decay functions for structural covariance matrices for surface area, cortical thickness and neurite density index (Fig. S2).

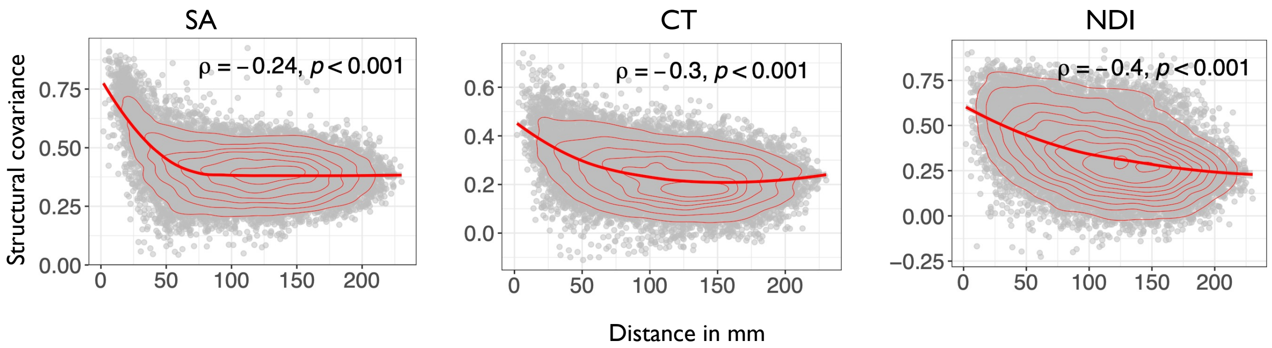

**Figure S2.** Distance decay function for surface area, cortical thickness and neurite denisty index based on structural covariance. Correlation (Spearman's) between structural covariance (y-axis) and geodesic distance in millimeters (x-axis).

#### 2.3 Enrichment of structural covariance and genetic similarity in Mesulam atlases

Structural covariance and genetic similarity showed similar relationships to classes of laminar differentiation. For CT structural covariance and genetic similarity were enriched in idiotypic and heteromodal class (Fig. S3). Genetic similarity of cortical thickness was higher in heteromodal ($z = 3.94$, $P < 0.05$) and idiotypic ($z = 2.85$, $P < 0.05$) classes. Structural covariance showed the same relationship cytoarchitectonic classes ( CT heteromodal $z = 3.46$, $P < 0.05$; idiotypic $z = 2.48$, $P < 0.05$).

#### 2.4 Enrichment of structural covariance and genetic similarity in Yeo networks

Structural covariance and genetic similarity showed similar relationships to Yeo networks. For SA structural covariance and genetic similarity were enriched in sensory-motor and default mode networks. Additionally genetic similarity for SA was enriched in dorsal attention networks and CT in fronto-parietal networks (Fig. S4). For surface area, genetic similarity was higher among regions within the sensory-motor ($z = 3.24$, $P < 0.05$) and default mode ($z = 2.84$, $P < 0.05$) networks relative to random networks. Structural covariance showed the same relationship to networks (SA sensory-motor $z = 3.03$, $P < 0.05$; default mode $z = 2.6$, $P < 0.05$).

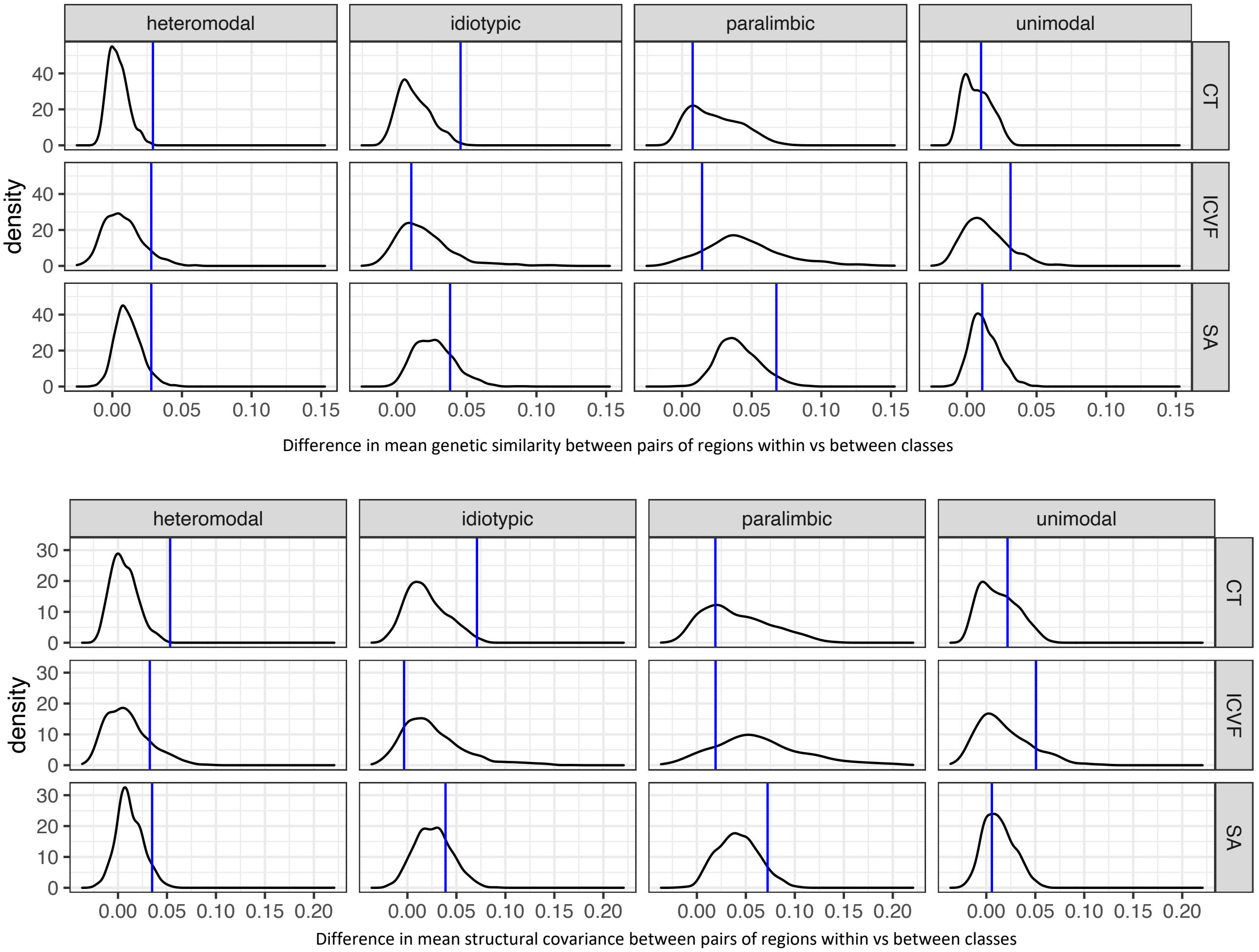

**Figure S3.** Enrichments in known cortical atlases (Mesulam classes). Specifically, we tested whether genetic similarity or structural covariance were higher or lower than expected by chance within regions of the same class compared to regions between classes. Significance was assessed by spin-permutation. Shown are permutation results from enrichments tests for mesulam classes. Blue lines indicate true value.

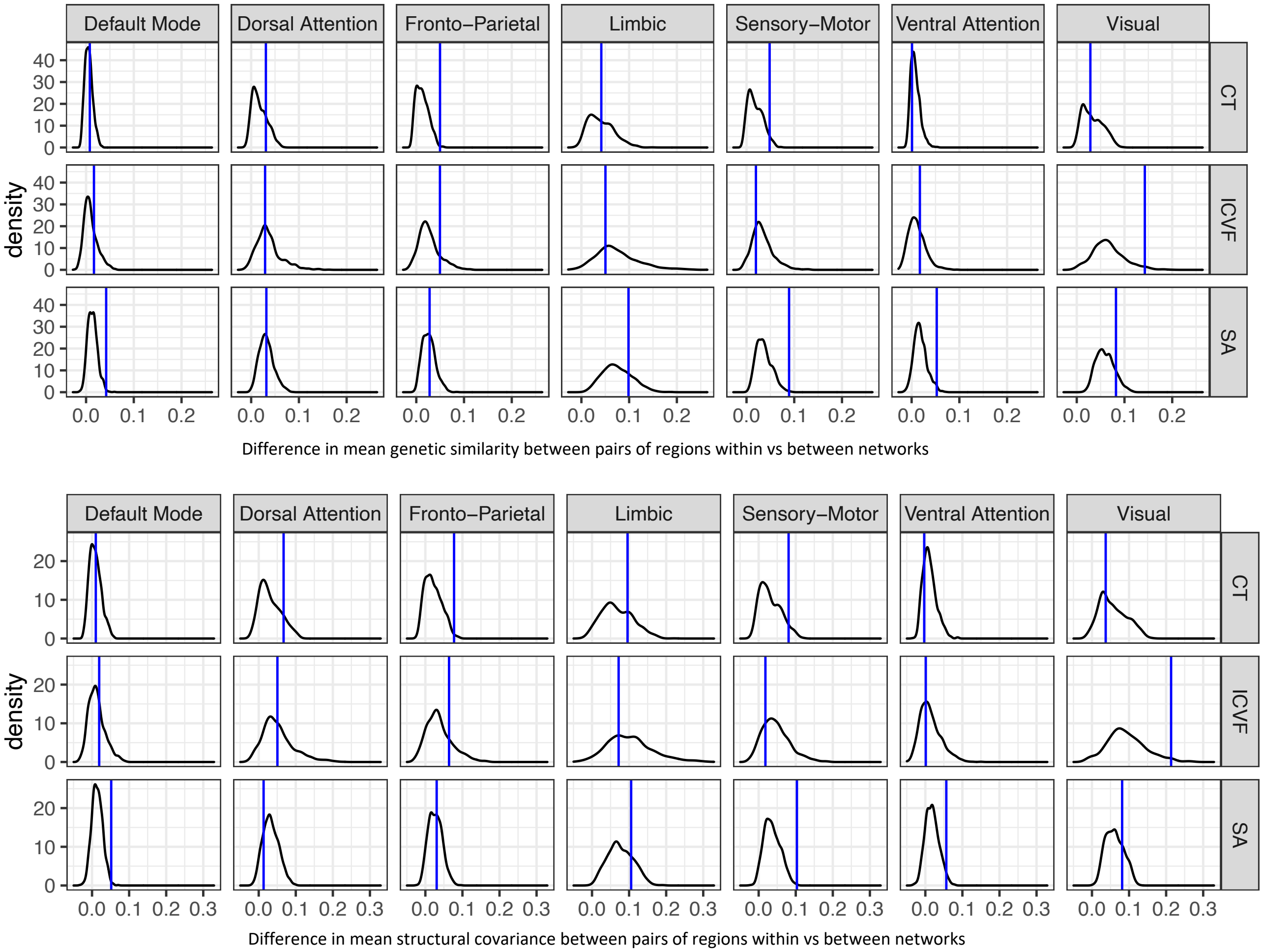

**Figure S4.** Enrichments in known cortical atlases (Yeo networks). Specifically, we tested whether genetic similarity or structural covariance were higher or lower than expected by chance within regions of the same network compared to regions between networks. Significance was assessed by spin-permutation. Shown are permutation results from enrichments tests for yeo networks. Blue lines indicate true value.

#### 2.5 PLS weights and intra- & inter-modular degree

We tested whether PLS1 weights were significantly correlated with intra- and inter-modular degree using spearman correlations. Inter and intra-modular degree were based on structural covariance matrices. We found significant positive correlations for all MRI metrics (Fig. S5).

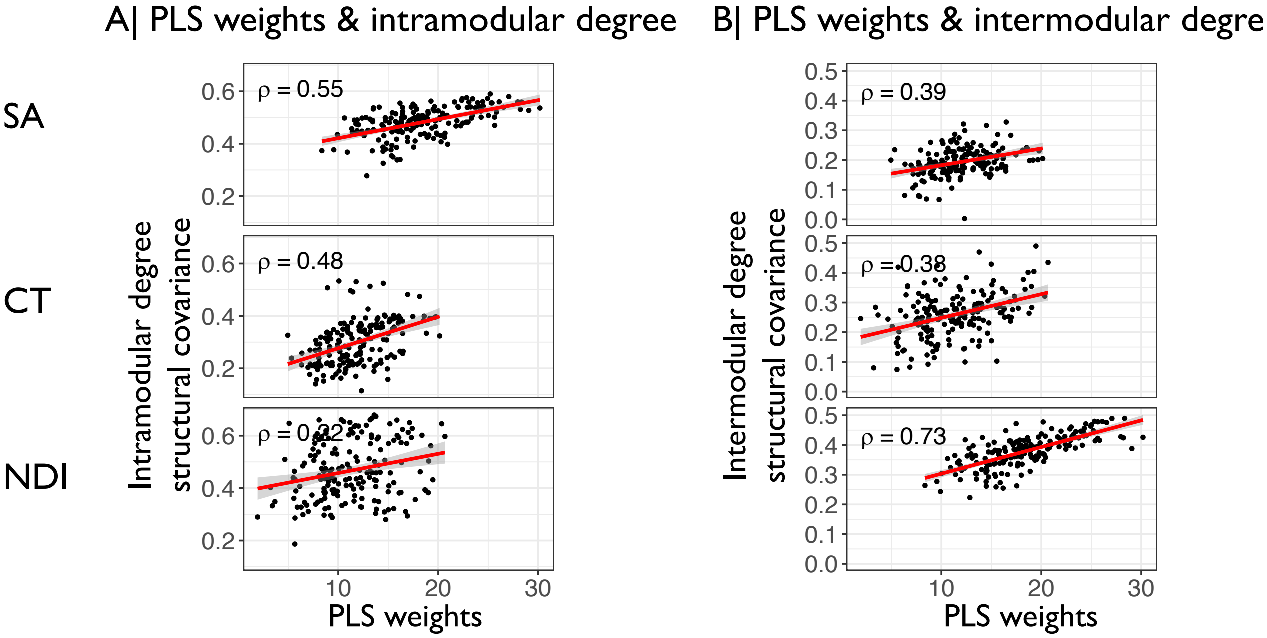

**Figure S5.** Show are correlations between PLS weights and intra (A) and inter-modular degree (B) for surface area, neurite denisty index and cortical thickness.

#### 2.6 Enrichment for constrained genes

We tested whether genes covarying between schizophrenia and MRI metrics were enriched for constrained genes. The analysis was performed on two inclusion thresholds. We found significant enrichments for all metrics at both thresholds (Fig. S6A-B).

#### 2.7 Cell type specific enrichment

Results of cell type specific enrichment analysis of covarying genes. Results are shown for both inclusion thresholds (Fig. S7A-B).

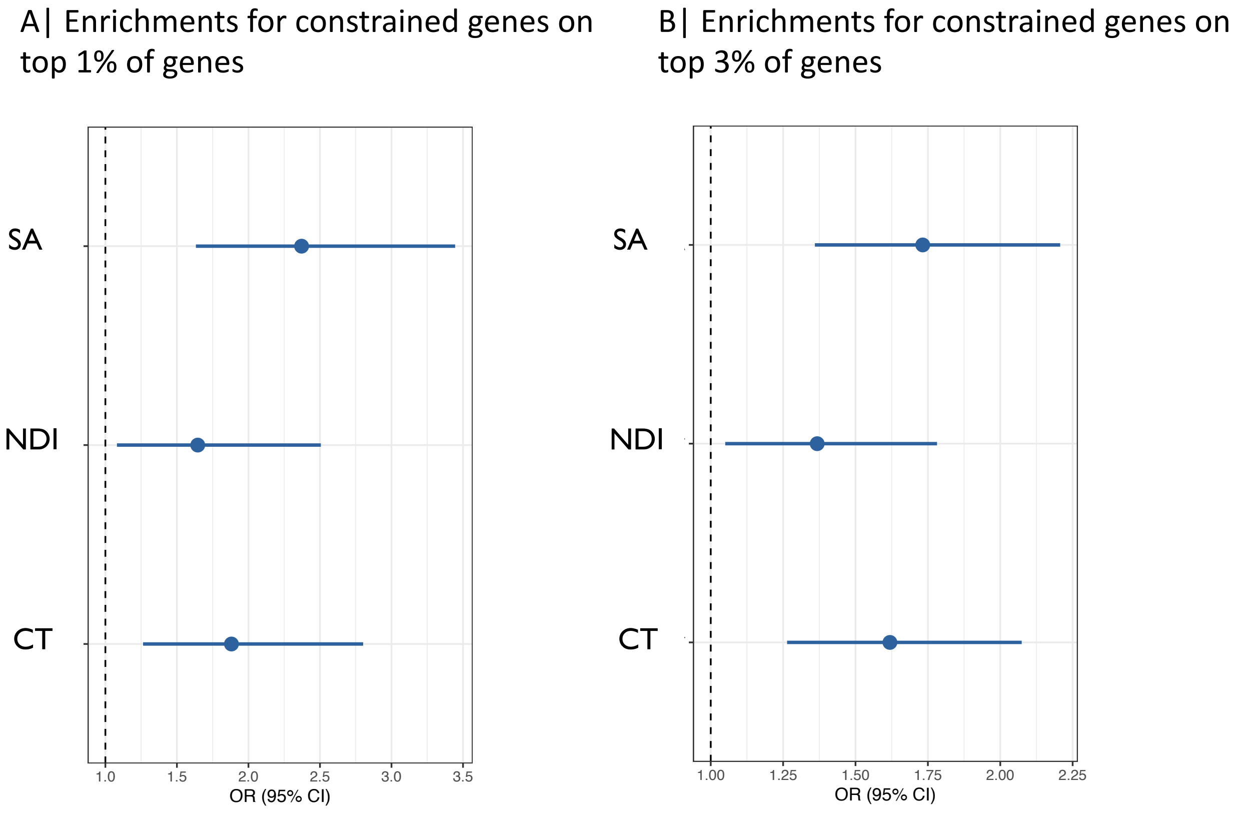

**Figure S6.** Enrichment results for constrained genes for the top 1% (A) and 3% (B) of genes with the highest influence on the covariation between brain structure and schizophrenia for surface area, neurite denisty index and cortical thickness.

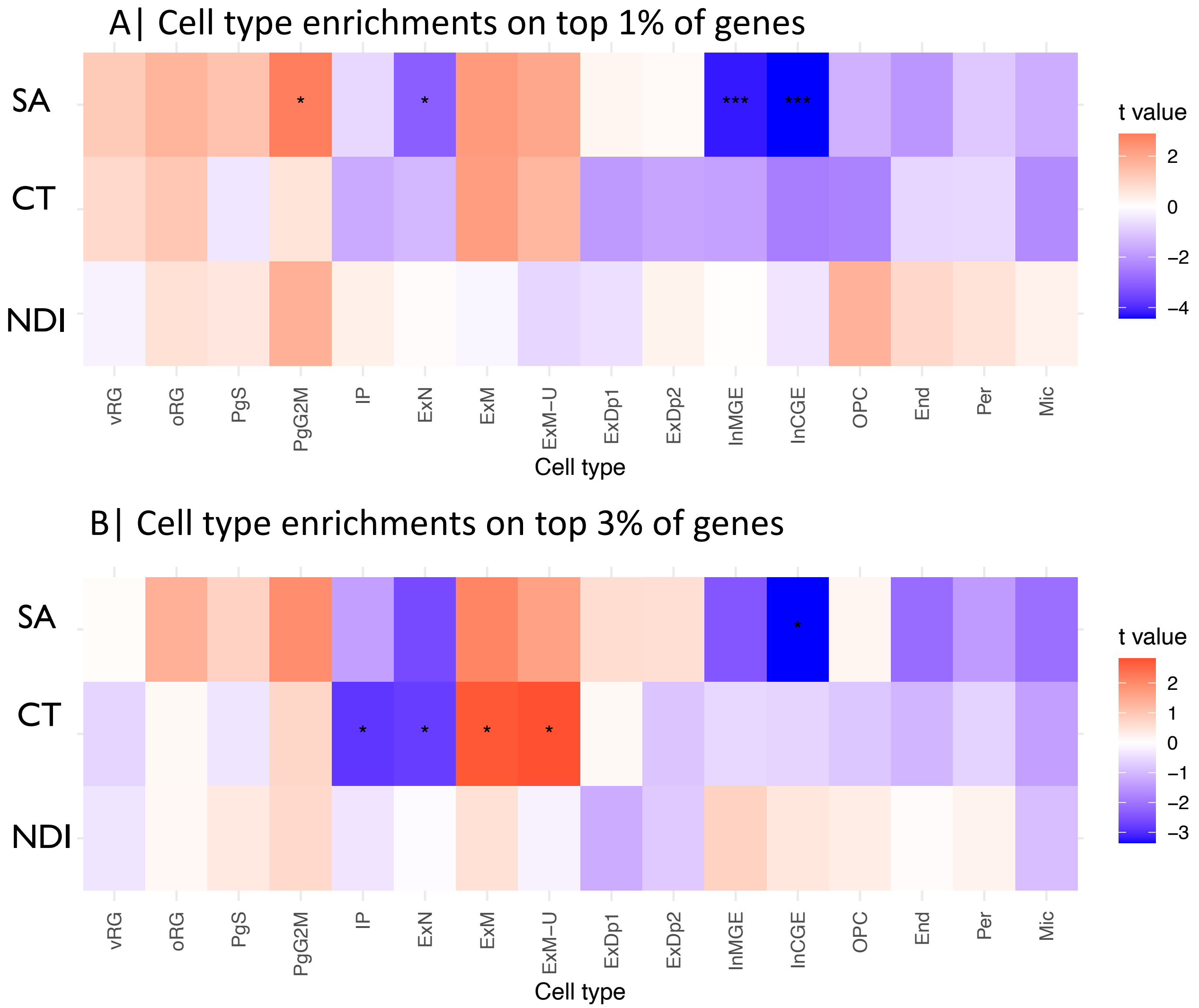

**Figure S7.** Cell type enrichments of the top 1% (A) and 3% (B) of genes with the highest influence on the covariation between brain structure and schizophrenia. Cell types: vRG = ventral radial glia, oRG = outer radial glia, PgS and PgG2M = cycling progenitors, S phase and G2-M phase respectively, IP = intermediate progenitors, ExN = Migrating excitatory neurons, ExM = Maturing Excitatory neurons, ExMU = Maturing excitatory neuron, upper enriched, ExDp1 = Excitatory deep layer neurons 1, ExDp2 = Excitatory deep layer neurons 2, InMGE = MGE Interneuron, InCGE = CGE interneuron, OPC = oligodendrocyte precursor cells, End = endothelial cells, Per = pericytes.
